# Comprehensive Immunophenotyping of Monocytes and Dendritic Cells Suggests Distinct Pathophysiology in Chronic Fatigue Syndrome and Long COVID

**DOI:** 10.64898/2026.04.10.26350613

**Authors:** Steliyan Petrov, Martina Bozhkova, Mariya Ivanovska, Teodora Kalfova, Dobrina Dudova, Yana Todorova, Radostina Dimitrova, Mariana Murdjeva, Hristo Taskov, Maria Nikolova, Michael Maes

## Abstract

Myalgic encephalomyelitis/chronic fatigue syndrome (ME/CFS) and long COVID are complex chronic conditions that often follow infectious triggers with overlapping clinical features but poorly defined pathophysiological relationships. This study aimed to identify disease-specific immune signatures through multiparameter immunophenotyping of monocytes, dendritic cells, and T-cell subsets. A total of 207 participants were included (ME/CFS: n = 103; long COVID: n = 63; healthy controls: n = 41). Peripheral blood mononuclear cells were analyzed using multiparameter flow cytometry. Statistical analyses included non-parametric testing, age-adjusted ANCOVA, correlation network analysis, and principal component analysis (PCA). Long COVID was characterized by increased M2-like monocyte polarization, elevated CD80 expression across monocyte subsets, expansion of dendritic cells, and reduced expression of activation markers, indicating persistent immune activation with features of immune exhaustion. In contrast, ME/CFS exhibited reduced costimulatory molecule expression, impaired CCR7-mediated immune cell trafficking, and less coordinated activation patterns, consistent with a state of immune suppression. Correlation network analysis revealed more extensive and integrated immune interactions in long COVID, while PCA identified distinct immunophenotypic components and enabled moderate discrimination between the two conditions. These findings demonstrate that ME/CFS and long COVID are characterized by distinct immune profiles, supporting the concept of divergent immunopathological mechanisms. The identified signatures may contribute to biomarker development and guide targeted therapeutic approaches.

## 1. Introduction

Myalgic encephalomyelitis/chronic fatigue syndrome (ME/CFS) and long COVID are debilitating complex chronic conditions that often follow infectious triggers. They have emerged as significant public health challenges, affecting millions of individuals worldwide [1,2]. Both conditions share striking clinical similarities, including intractable fatigue, post-exertional malaise, cognitive dysfunction ("brain fog"), and dysautonomia with orthostatic intolerance [1,3,4]. The COVID-19 pandemic has dramatically increased awareness of post-viral syndromes, with approximately 10% of SARS-CoV-2-infected individuals developing long COVID, and a substantial proportion of patients meeting the diagnostic criteria for ME/CFS [5,6].

The phenotypic similarities between ME/CFS and long COVID encompass multiple pathophysiological domains. Both conditions demonstrate autonomic dysfunction, with comparable rates of reduced orthostatic cerebral blood flow velocity, widespread autonomic failure, small fiber neuropathy, and postural orthostatic tachycardia syndrome [1,7–10]. It was hypothesized that the somatic and mental symptoms of both long COVID and CFS are underpinned by activated immune-inflammatory and oxidative and nitrosative stress (IO&NS) pathways, as a result of imbalanced immune regulatory mechanism [11,12]. Indeed, at the molecular level, both conditions exhibit elevated oxidative stress in peripheral blood lymphocytes, with aberrations in reactive oxygen species clearance pathways, elevated glutathione levels, and mitochondrial dysfunction [2]. Proteomic analyses of peripheral blood mononuclear cells have identified overlapping protein clusters and enriched molecular pathways, particularly involved in immune functions and mitochondrial energy production [5]. Several studies have tried to elucidate immune phenotypic and functional signatures describing this imbalance and possibly differentiating between the two conditions.

Standard laboratory tests and routine clinical assessments consistently fail to distinguish between ME/CFS and long COVID. Novak et al. demonstrated that comprehensive laboratory analyses covering metabolic, inflammatory, autoimmune, and hormonal profiles did not differentiate between the two conditions [1]. Similarly, Kedor et al. found that disease severity and symptom burden were comparable between post-COVID-19 ME/CFS and non-COVID-19 ME/CFS patients, with standard biomarkers showing similar patterns [13]. This diagnostic limitation has significant clinical implications, as both conditions currently lack approved treatments and rely on symptom-based diagnostic criteria [2,14]. The absence of distinguishing biomarkers has fuelled debate about whether long COVID and ME/CFS represent the same condition with different triggers or fundamentally distinct pathophysiological entities requiring different therapeutic approaches.

Emerging evidence suggests that deeper immunological investigation may reveal important distinctions obscured by conventional testing. Gene expression analyses using immune exhaustion panels have demonstrated that ME/CFS is characterized by downregulated interferon signalling and immunoglobulin genes, suggesting a state of immune suppression or exhaustion, while long COVID exhibits dysregulated expression of genes related to antigen presentation, cytokine signalling, and persistent immune activation [15]. Single-cell RNA sequencing has revealed extensive peripheral immune remodelling in long COVID with ME/CFS features, including marked reduction in naïve T cells, regulatory T cells, MAIT cells, and γδT cells, accompanied by monocyte transcriptional skewing—alterations that are less pronounced in idiopathic ME/CFS [16].

Understanding how different immune cell populations and activation states differ between these conditions may provide insights into the mechanisms driving chronic immune dysregulation. Monocytes and monocyte-derived dendritic cells are strategic immune components that participate all along the inducing, effector, and regulatory stages of adaptive immunity, and may contribute to its imbalance. While M1- like monocytes are activated by pathogen invasion and primarily promote inflammation and tissue damage, the M2 –like subset, on the other hand, have anti-inflammatory properties, and are involved in angiogenesis, tissue repair and remodelling [17]. DCs are both involved in innate defence and the initiation and regulation of adaptive T-cell responses [18]. As monocytes and dendritic cells play crucial roles in the immune response to COVID-19, their dysfunction can contribute to immune imbalance underlying the persistence of symptoms [19]. Thus, long-term changes observed in the levels of certain monocyte and dendritic cell subsets, as well as the extensive epigenetic changes in monocytes from convalescent patients, exposing genes related to cytokine production and leukocyte activation were proposed to enhance chronic inflammation and tissue damage observed in long COVID [20,21]. Studies on monocyte subsets in chronic fatigue syndrome (CFS) also revealed significant changes in the balance between M1 and M2 subsets, and the expression of some monocyte specific markers [22]. However, comparative comprehensive characterization of these compartments in long Covid and ME/CFS are lacking.

High-dimensional flow cytometry methods enabling simultaneous measurement of over 40 markers on individual cells provide comprehensive assessment of immune cell subsets and their functional states [23]. Such approaches have identified unique immunotypes within ME/CFS based on cerebrospinal fluid immune phenotyping, revealing distinct patient clusters with different pathogen exposure histories and inflammatory profiles despite similar clinical presentations [24]. Traditional flow cytometry studies (typically measuring 6-12 markers) have identified various immune alterations in ME/CFS, including changes in NK cell subsets, T cell populations, and dendritic cell function, but these have not consistently identified distinct immunotype clusters [25–27]. In long COVID, multidimensional immune profiling has identified distinguishing features including marked differences in circulating myeloid and lymphocyte populations, exaggerated humoral responses against SARS-CoV-2, elevated antibody responses against non-SARS-CoV-2 viral pathogens (particularly Epstein-Barr virus), and reduced cortisol levels [28]. Combining biomarkers of endothelial dysfunction and inflammation with clinical outcome measures has enabled differentiation between ME/CFS and long COVID using discriminant analysis of principal components, demonstrating that multiparametric approaches can achieve diagnostic separation where individual markers fail.

The present study addresses further this critical knowledge gap by performing multiparameter immunophenotyping of monocyte subsets (M1-like and M2-like), dendritic cells, and T cell populations in patients with ME/CFS, and long COVID, in relation to the severity of psychosomatic symptoms, and healthy controls. We hypothesized that the implementation of correlation network analysis and principal component analysis might help identify disease-specific immune signatures that could serve as composite biomarkers and provide mechanistic insights into the distinct pathophysiology of ME/CFS and long COVID.

## 2. Results

### 2.1. Participants

A total of 207 participants were included in the study. The demographic characteristics of the subgroups are given in Table 1. The overall cohort included 54 males and 153 females. The sex distribution did not differ significantly between the study groups (χ² = 0.454, df = 2, p = 0.797). As expected, a higher proportion of females was observed in the CFS group, consistent with the known female predominance of this condition reported in epidemiological studies. Age differed significantly between healthy subjects and the other two groups (Kruskal–Wallis H = 24.79, df = 2, p < 0.001), indicating heterogeneity in age distribution among healthy controls, CFS patients, and LC individuals (Table 1).

**Table 1.** Demographic characteristics of the study population.

| Variable | HC (n=41) | CFS (n=103) | LC (n=63) | p value |
| --- | --- | --- | --- | --- |
| Male, n(%) | 9 (22.0) | 28 (27.2) | 17 (27.0) | 0.797 |
| Female, n(%) | 32 (78.0) | 75 (72.8) | 46 (73.0) |  |
| Age, median (IQR) | 27 (22) | 48 (20) | 48 (19) | <0.001 |

Participants with CFS had a longer median duration since symptom onset (36 months) compared to those with long COVID (23 months, p=0.001), with sex-specific medians of 24 and 36 months in men and women with CFS, and 25 and 20 months in men and women with long COVID, respectively.

### 2.2. M2- like monocytes

The distribution of monocyte polarization subsets differed between groups. The proportion of M2-like monocytes increased in chronic fatigue syndrome patients (median 0.10%), and even more so in LC patients (median 0.20%), with a statistically significant increase in LC as compared to HC (median 0.1%) (fig.1-A). Analysis of activation markers (fig.1-B) showed that the MFI of CD69⁺CD38⁺ M2-like monocytes was significantly lower in LC (median 135155), while comparable levels were observed between HC (median 193425) and CFS (median 193851). The expression of the costimulatory molecule CD80 showed changes between the groups. Specifically, M2-like CD80 MFI was highest in LC (median 230), statistically exceeding CFS (median 137.1), while HC showed intermediate levels (median 143.4) (fig.1-C). CD197 expression was lowest in CFS (MFI 345), as compared to LC (408) and HC (458), without reaching statistical significance (data not shown).

**Figure 1.**
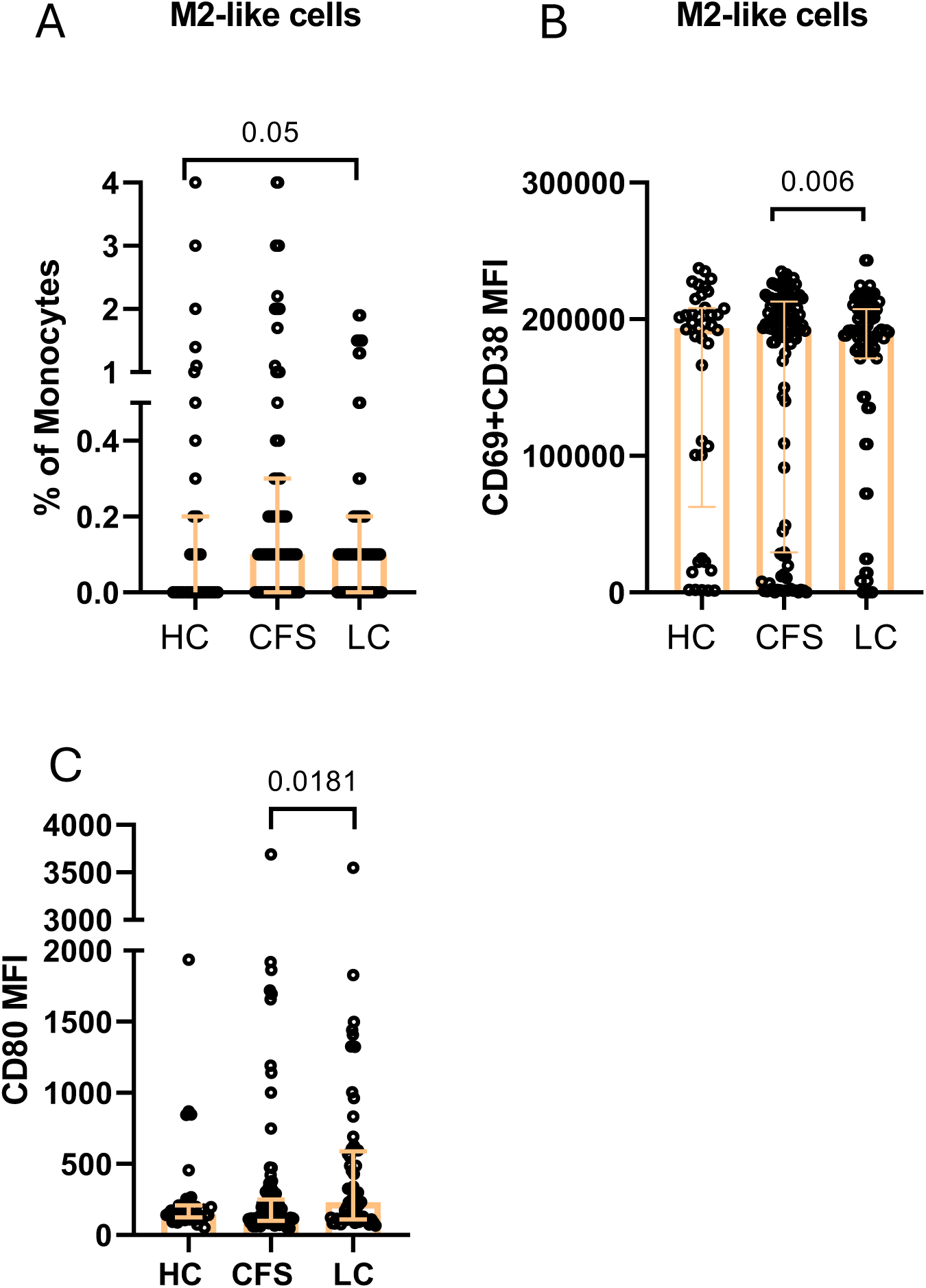
Immunophenotypic characterization of M2-like cells: A) Distribution in the three studied groups: B) CD69+CD38 expression (MFI) C) CD80 expression (MFI). Individual values are presented (circles), together with median values and interquartile ranges (whiskers) ; Exact *p* values (Age-adjusted ANCOVA test results are provided).

### 2.3. M1-like monocytes

A similar trend was observed for M1-like monocytes in terms of CD80 MFI, which was increased in LC (median 157.8), as compared to HC (median 118.4), and CFS (median 112.2, p<0.05) (fig.2-A). Chemokine receptor expression also differed between groups. LC and CFS were distinguished with a significantly decreased CD197 (CCR7) expression on M1-like cells: (median MFI) 236 and 280 for CFS and LC, vs. 315 for HC (fig.2-B). Likewise, CD69+CD38 expression decreased in CFS (11841), and LC (10249) as compared to HC (12508), without reaching statistical significance.

**Figure 2.**
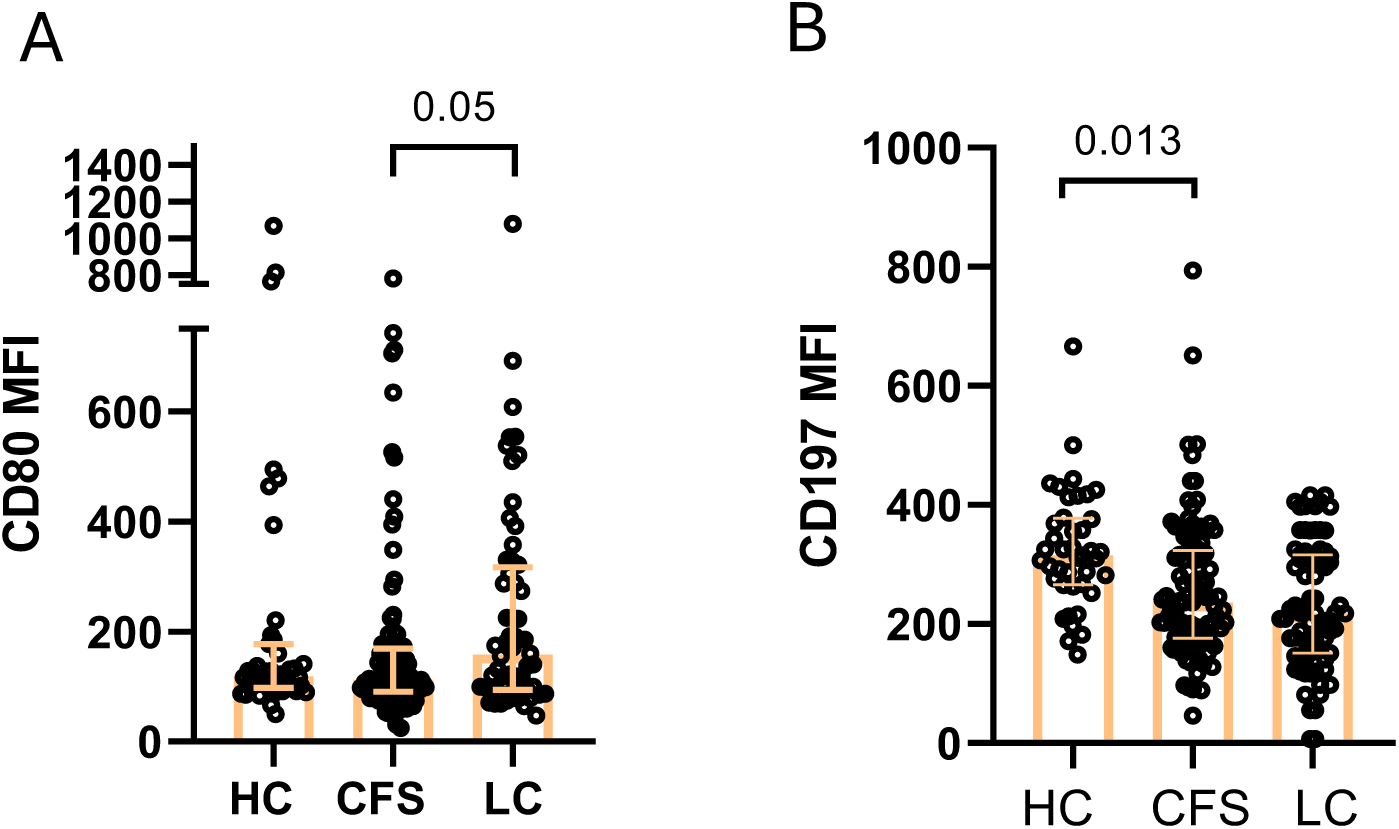
Immunophenotypic characterization of M1-like cells in the three studied groups: A) CD80 expression (MFI) B) CD197 expression (MFI, η²ₚ = 0.004). Individual values are presented, together with median values and interquartile ranges; Exact *p* values (Age-adjusted ANCOVA) are provided.

### 3.4. Dendritic cells

The frequency of circulating dendritic cells also differed across the study groups (fig.3-A). The highest levels were detected in LC patients (median 9.7%), significantly exceeding CFS (median 7%) and HC (median 7.5%). Activation marker analysis revealed that the expression of activation markers on dendritic cells was lower in CFS as compared to LC (CD38+CD69+ MFI) 2941 vs. 5532. Likewise, CD80 expression was lowest in CFS (MFI) 380 vs. 438 and 483 for LC and HC. (fig.3-B). A similar trend was observed for CD197 expression which was decreased in LC (median MFI 263) and CFS (266.5) as compared to HC (308.5), data not shown.

**Figure 3.**
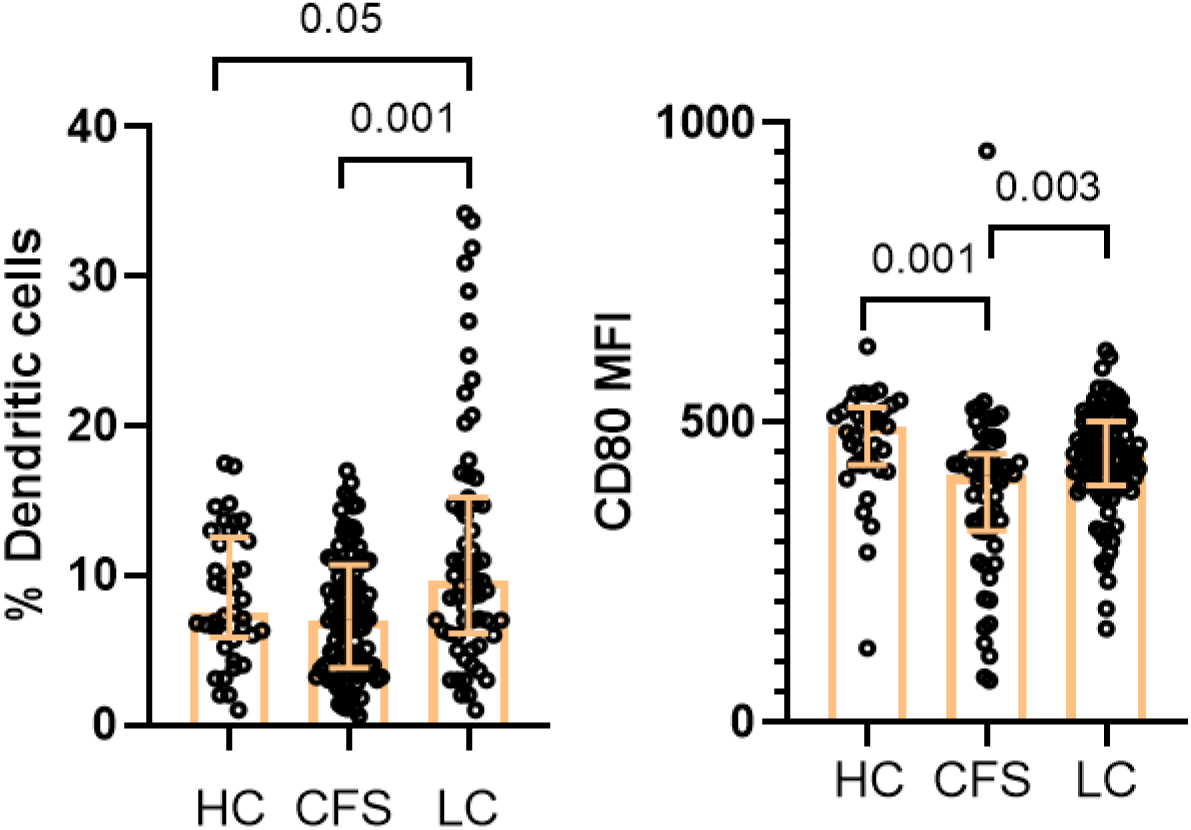
Immunophenotypic characterization of DC in the three studied groups: A) Total DCin the three studied groups; B) CD69+CD38 (MFI). Individual values are presented (circles), together with median values and interquartile ranges (whiskers); Exact p values (Age-adjusted ANCOVA) are provided.

### 3.5. Symptoms severity analysis

A composite score was calculated based on the total number of points from the three questionnaires (HAM-A, HAM-D, FibroFatigue). Exploratory threshold was set at 50. People with score above the threshold were considered to have moderate symptomatology, and those with a total score under 50 were considered as mild cases. Both CFS (median 51.50) and LC (median 48.00) had significantly higher total scores, compared to HC (median 25.00)(fig.4).

**Figure 4.**
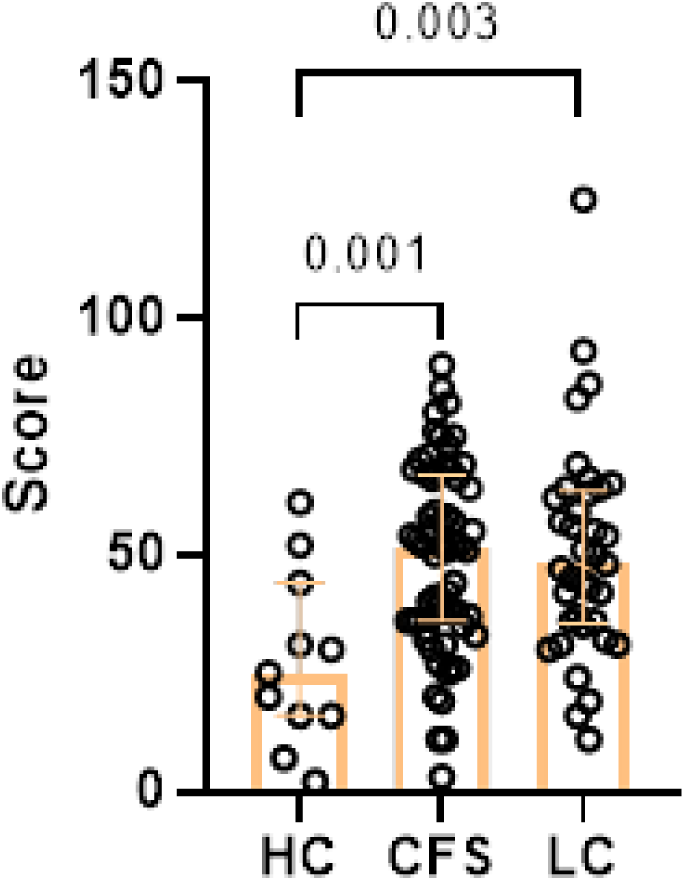
Results from age-adjusted ANCOVA, showing total score from the three questionnaires in the three studied groups as individual values (circles). Exact *p* values are provided. Results are presented as median values and interquartile ranges (whiskers).

Comparison of mild and moderate disease forms did not affect the interactions between M1-like/M-2 like parameters, but revealed significant differences in T-cell phenotypes in terms of surface expression of CD45-RA and CD62L. Patients with moderate disease exhibited significantly higher percentages of CD3⁺CD4⁺CD62L⁺ T cells (median for CFS 16.20%, median for LC 17.0%) compared with HC (median 11.3%) (fig.5-A). Conversely, the proportion of CD3⁺CD4⁺CD45RA⁺ T cells was significantly lower in moderate CFS and LC (medians CFS 20.05%, and LC 17.7% respectively) as compared to HC (median 24.1%) (fig.5-B).

**Figure 5.**
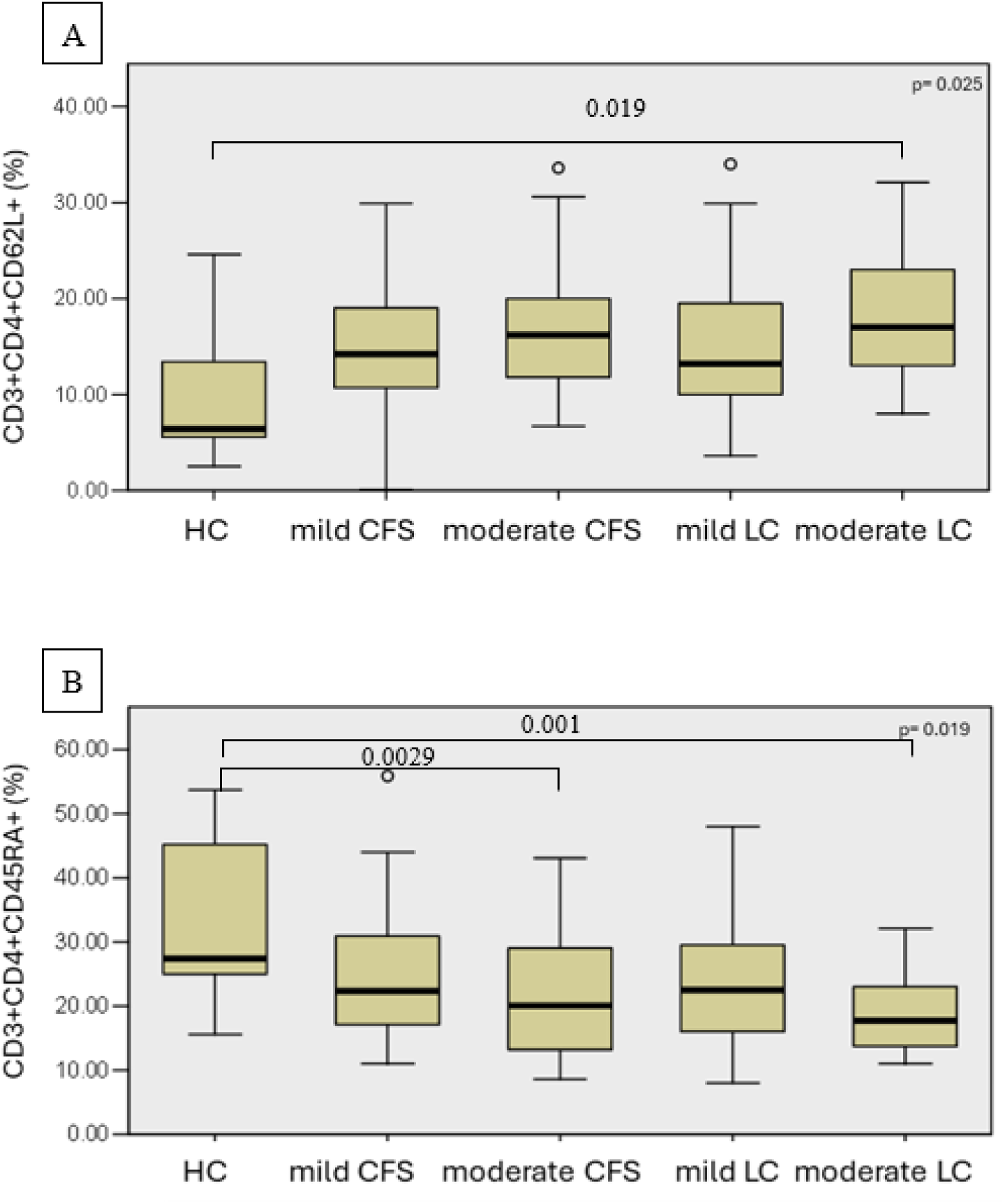
Еffect of symptoms severity on T-cell differentiation status in the three studied groups. A) Percentage of CD3+CD4+CD62L+ T cells, B) Percentage of CD3+CD4+CD45RA+ T cells. Results are presented as median values and interquartile ranges. Exact *p* values from age-adjusted ANCOVA are provided.

Additionally, patients with moderate LC showed significantly increased expression of CD69⁺CD38 activation markers on their DC as compared to mild cases and HC (median 6012 vs. 1259,5 and 3906). At the same time, CD80 expression was decreased on DC from moderate LC cases (median 89, 3) as compared to mild cases and HC (median 222,5 and 95,53 respectively). The patients with moderate CFS also showed significantly increased MFI of CD69⁺CD38⁺ dendritic cells (median 5486) and decreased CD80 expression (median 79,9) as compared to mild cases, and HC (84,10 and 95,53) (fig.6).

**Figure 6.**
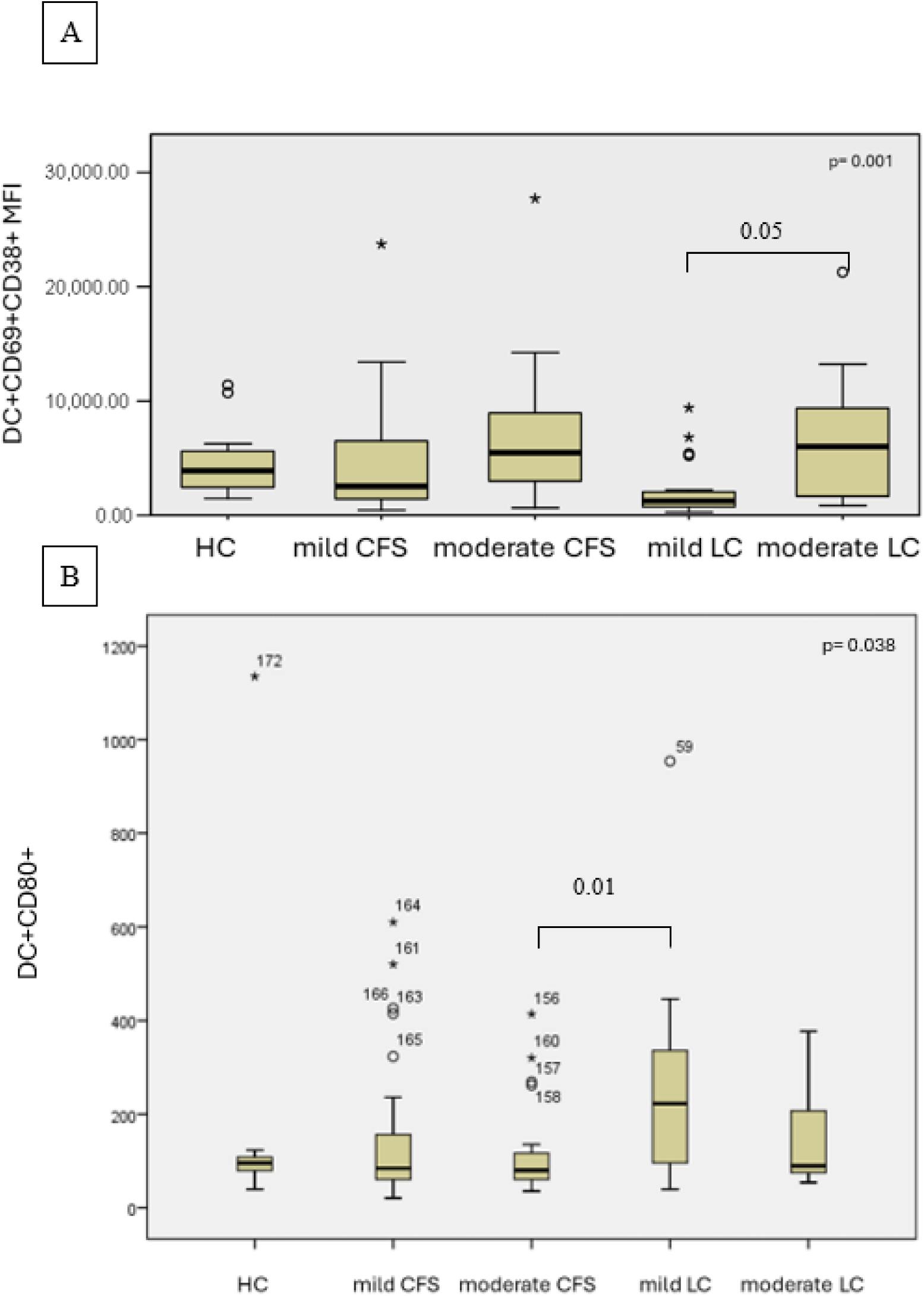
Еffect of symptoms severity on DC functional status in the three studied groups. A) Percentage of CD69+CD38+ expression (MFI), B) CD80 expression (MFI). Results are presented as median values and interquartile ranges. Exact p values from age-adjusted ANCOVA are provided.

### 2.5. Correlation analysis

In CFS patients, strong positive correlations were observed between several activation markers within the monocyte compartment (fig.7-A). Notably, HLA-DR expression correlated between M2-like and M1-like monocytes, and CD80 expression showed a positive correlations with HLA-DR in both M1-like and M2-like populations. Additionally, CD80 expression of M1-like monocytes correlated positively with the total number of M2-like monocytes. Within M2-like cells, CD80 expression positively correlated with HLA-DR and CD197. Similarly, CD80 and CD197 expressions in M1-like cells showed a strong positive association. A strong positive correlation was also observed between CD95 expression on CD4⁺ and CD8⁺ T cells, as well as between naïve T-cell populations defined as CD62L⁺CD45RA⁺ within CD4⁺ and CD8⁺ T cells. Several strong negative correlations were detected. The total number of DC inversely correlated with total M1-like monocytes. In addition, CD80 expression within M1-like monocytes negatively correlated with the total M1-like cell numbers. Within M2-like cells, CD80 expression negatively correlated with CD206 and CD69+CD38 expression, while CD197 expression negatively correlated with CD206. Score from the pHAM-D correlated negatively with HLA-DR and CD80 on M2-like monocytes and HLA-DR on M1-like monocytes. Positive correlation was established with CD69+CD38 on M1-like monocytes.

**Figure 7.**
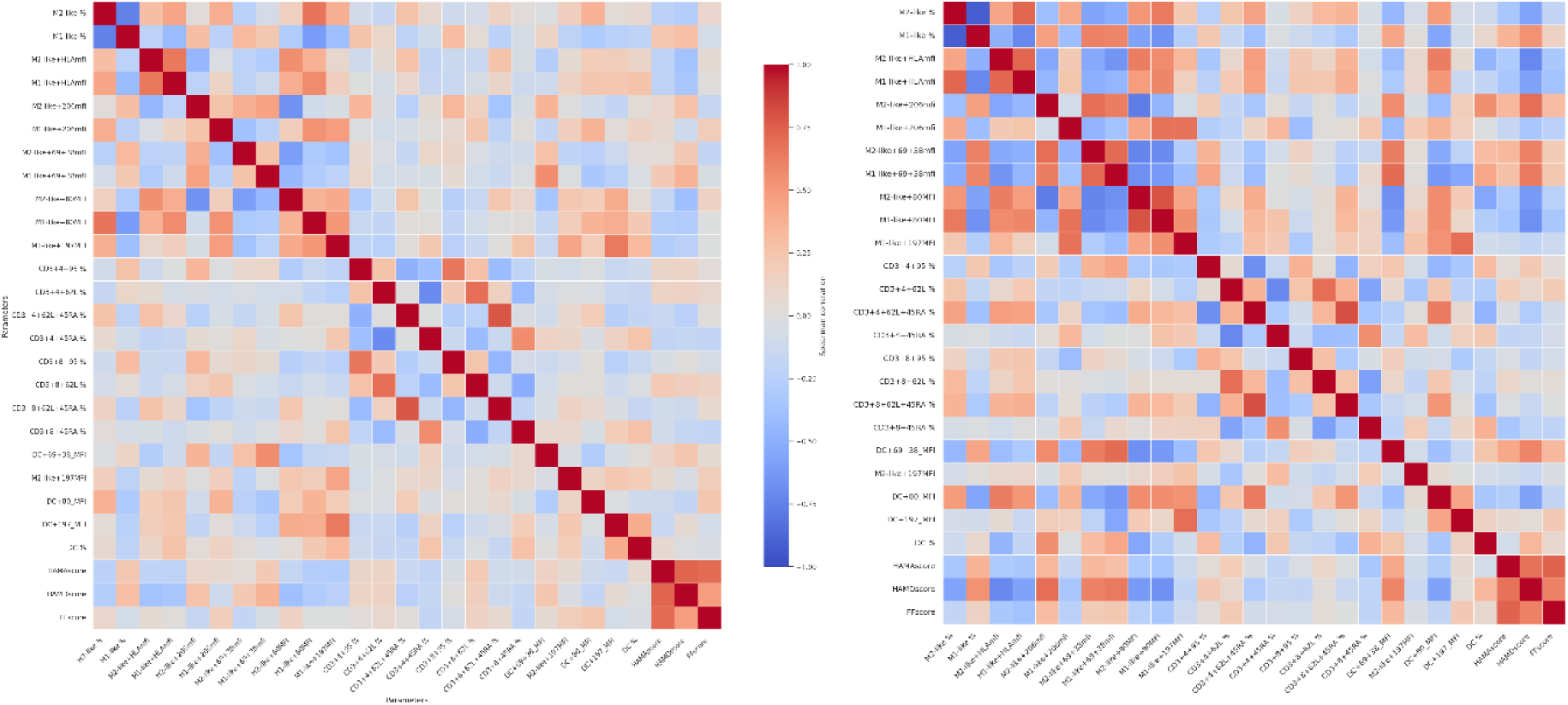
Heatmaps generated from a correlation matrix displaying pairwise correlations between the different phenotypic markers for CFS (A) and LC (B) samples. Colors indicate the strength and direction of the correlations (blue = negative, red = positive).Correlation coefficients were calculated using Spearman’s method. Significance levels were adjusted for multiple comparisons using the FDR correction. Relationships are shown between.

The correlation network in LC patients was more extensive and involved both monocyte subsets and dendritic cells (fig.7-B). Strong positive correlations were observed between HLA-DR expression on M1-like monocytes and total M2-like numbers, as well as between HLA-DR expression on M1-like and M2-like monocytes. Activation markers were also coordinated across subsets, with CD69+CD38 expression on M1-like cells positively correlating with the same markers on M2-like cells. In addition, M2-like CD69+CD38+ cells positively correlated with both M2-like CD206 expression and total M1-like numbers. Costimulatory signaling showed strong associations, with CD80 expression on M1-like cells positively correlating with HLA-DR, CD206, and CD197 expression across monocyte subsets. Similarly, CD197 expression was strongly correlated between M1-like and M2-like populations. DC were strongly integrated into this network. Total DC numbers positively correlated with total M2-like monocytes, HLA-DR expression on M1-like and M2-like cells, and CD80 expression across monocyte populations. Furthermore, CD69+CD38 DC expression correlated positively with activation markers (HLA-DR and CD80) in both M1-like and M2-like subsets. Several negative correlations were also detected in LC patients. Total M1-like monocytes inversely correlated with M1-like HLA-DR, M2-like CD80, and M2-like CD197 expression. Additionally, CD80 expression on M2-like cells negatively correlated with CD69+CD38 expression in both M1-like and M2-like populations. pHAM-D score correlated negatively with CD80 on DC and M1-like monocytes, HLA-DR on both M1 and M2-like monocytes. Positive correlations were reported with CD206 on M2-like monocytes, as well as with CD69+CD38 on both M1 and M2-like monocytes. pHAM-A scores correlated positively with CD69+CD38 on DC.

### 2.7. Principle component analysis

Principal component analysis without rotation was performed on the selected immune biomarkers across all participants to identify underlying dimensions of immune function. The Kaiser–Meyer–Olkin measure of sampling adequacy was 0.774, indicating good suitability for factor analysis. Bartlett’s test of sphericity was significant (p < .001), confirming that the correlation matrix was appropriate for PCA. PCA identified four components with eigenvalues greater than 1, which together explained a substantial proportion of the total variance (Table 2). The first component was characterized by high positive loadings (>0.6) of monocyte-associated markers, including HLA-related measures (M2-like+HLA-DR, M1-like+HLA-DR) and CD80 expression, alongside negative loadings of activation markers such as CD69/CD38 (Table 3). The second component showed strong loadings of T-cell differentiation markers, particularly CD4⁺ and CD8⁺ subsets reflecting naïve versus memory phenotypes (CD3+CD4+62L+CD45-RA-, CD3+CD4+CD45-RA+CD62L-, CD3+CD8+CD45-RA+CD62L-). The third component was defined by markers related to T-cell activation and maturation, including CD62L/CD45RA.

**Table 2.** Total variance explained by PCA.

| Component | Initial Eigenvalues |  |  | Extraction Sums of Squared Loadings |  |  |
| --- | --- | --- | --- | --- | --- | --- |
|  | Total | % of Variance | Cumulative % | Total | % of Variance | Cumulative % |
| 1 | 4.737 | 31.577 | 31.577 | 4.737 | 31.577 | 31.577 |
| 2 | 2.562 | 17.079 | 48.656 | 2.562 | 17.079 | 48.656 |
| 3 | 1.769 | 11.796 | 60.452 | 1.769 | 11.796 | 60.452 |
| 4 | 1.095 | 7.298 | 67.750 |  |  |  |
| 5 | .940 | 6.270 | 74.020 |  |  |  |
| 6 | .767 | 5.110 | 79.130 |  |  |  |
| 7 | .629 | 4.191 | 83.321 |  |  |  |
| 8 | .506 | 3.373 | 86.694 |  |  |  |
| 9 | .454 | 3.028 | 89.722 |  |  |  |
| 10 | .375 | 2.500 | 92.222 |  |  |  |
| 11 | .345 | 2.299 | 94.521 |  |  |  |
| 12 | .249 | 1.657 | 96.178 |  |  |  |
| 13 | .224 | 1.491 | 97.669 |  |  |  |
| 14 | .193 | 1.286 | 98.956 |  |  |  |
| 15 | .157 | 1.044 | 100.000 |  |  |  |

Extraction Method: Principal Component Analysis. PCA of selected immune biomarkers identified four components with eigenvalues greater than 1, accounting for a cumulative 67.75% of the total variance. The first component explained 31.57% of the variance, followed by the second (17.08%), and third (11.79%) components.

**Table 3.**
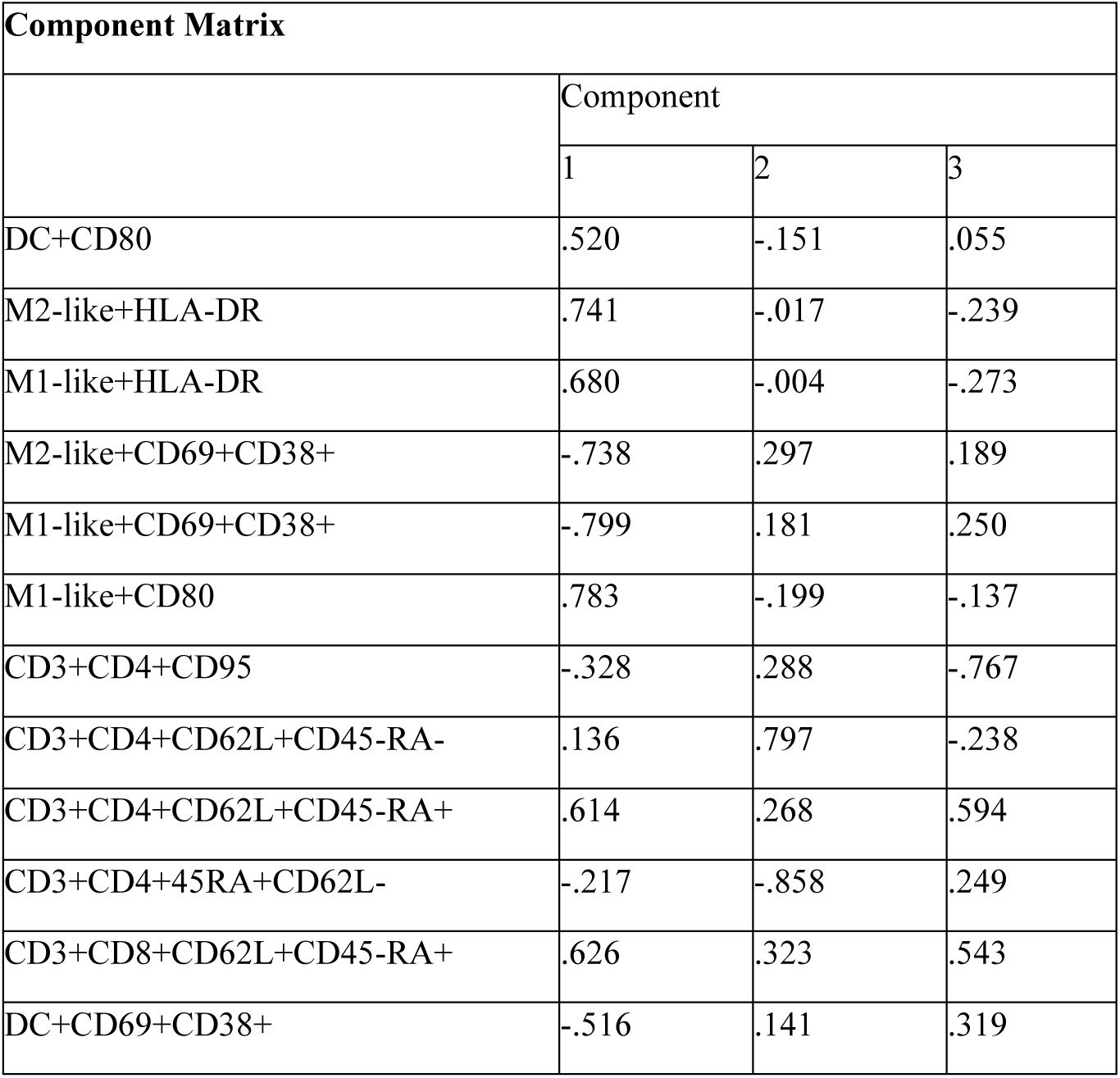
Component matrix showing loadings of immune biomarkers on principal components.

| Component Matrix |  |  |  |
| --- | --- | --- | --- |
|  | Component |  |  |
|  | 1 | 2 | 3 |
| DC+CD80 | .520 | -.151 | .055 |
| M2-like+HLA-DR | .741 | -.017 | -.239 |
| M1-like+HLA-DR | .680 | -.004 | -.273 |
| M2-like+CD69+CD38+ | -.738 | .297 | .189 |
| M1-like+CD69+CD38+ | -.799 | .181 | .250 |
| M1-like+CD80 | .783 | -.199 | -.137 |
| CD3+CD4+CD95 | -.328 | .288 | -.767 |
| CD3+CD4+CD62L+CD45-RA- | .136 | .797 | -.238 |
| CD3+CD4+CD62L+CD45-RA+ | .614 | .268 | .594 |
| CD3+CD4+45RA+CD62L- | -.217 | -.858 | .249 |
| CD3+CD8+CD62L+CD45-RA+ | .626 | .323 | .543 |
| DC+CD69+CD38+ | -.516 | .141 | .319 |

### 3.8. Associations Between Immune Components and Clinical Symptoms

Multiple regression analysis was performed to examine the association between PCA-derived immune components and anxiety symptoms (HAM-A), controlling for age and sex.

The model revealed that Component 1 (innate immune activation axis) was significantly and inversely associated with anxiety (β = -0.23, p = .015), while Component 2 (T-cell differentiation axis) was positively associated with anxiety (β = 0.20, p = .035). Component 3, age, and sex were not significant predictors.

A similar regression model was conducted for depressive symptoms (HAM-D).

Component 1 showed a strong negative association with depression (β = -0.50, p < .001), whereas Component 2 was positively associated with depression severity (β = 0.31, p < .001). Additionally, sex was a significant predictor (β = 0.20, p = .011), while age and Component 3 were not significant.

For fatigue (FF), the overall regression model was not statistically significant. However, Component 2 (T-cell differentiation axis) remained a significant positive predictor of fatigue severity (β = 0.26, p = .007). No significant associations were observed for Component 1, Component 3, age, or sex.

Partial least squares discriminant analysis was performed on the selected immune biomarkers in the clinically more homogeneous subgroup of participants with total questionnaires score above 50 (fig.8-A).

**Figure 8.**
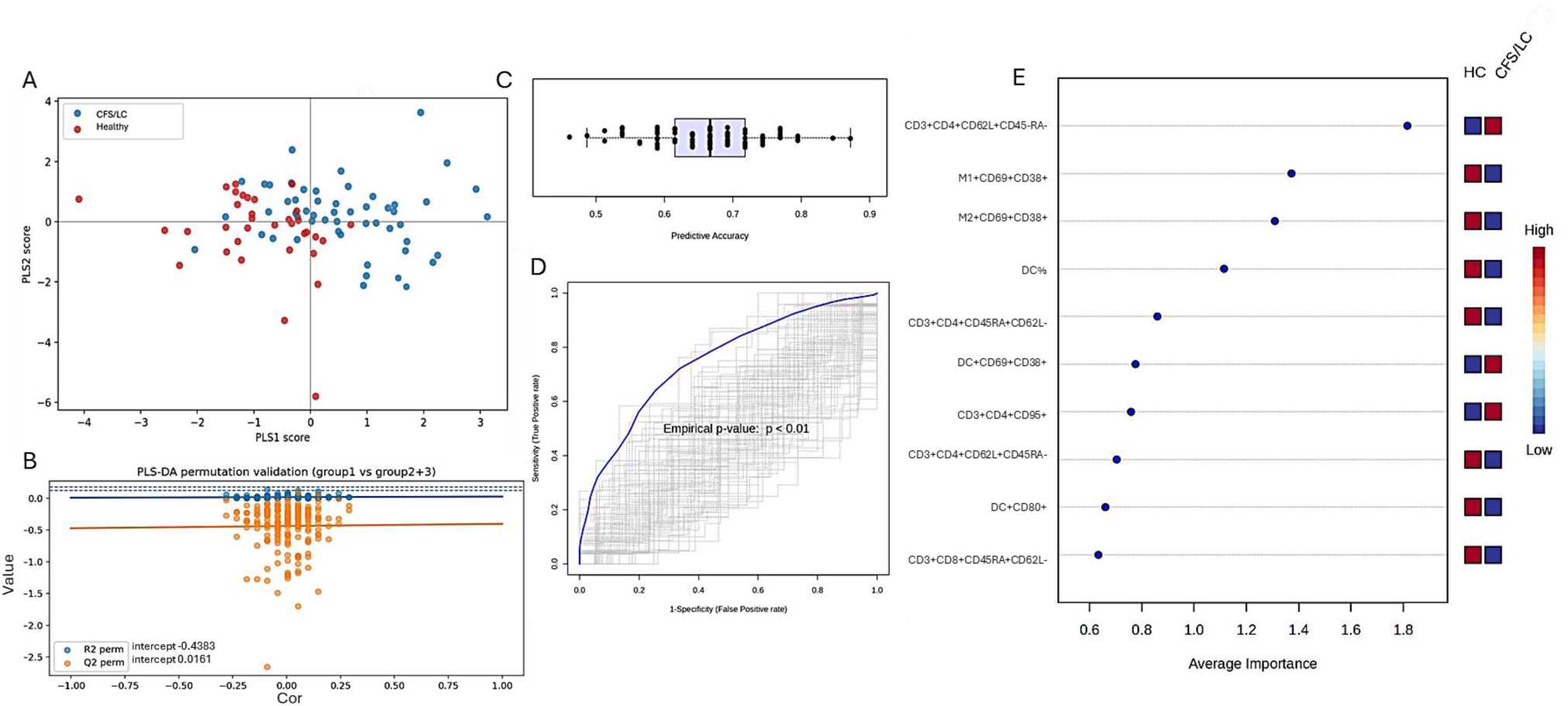
A) Scatter plot of predicted class probabilities for individual samples. Each point represents a participant, with colors indicating group membership (HC vs LC/CFS); B) Permutation test for model validation. The original model (blue line) demonstrates higher predictive performance compared to permuted models (orange points), with intercept values indicating that the model is not overfitted (empirical p < 0.01); C) Cross-validated predictive accuracy across different numbers of features. Model performance improves with increasing feature number, reaching an accuracy of approximately 69%, indicating moderate classification capability. D) ROC curve for the PLS-DA model. The model achieved an area under the curve (AUC) of 0.758 (95% CI: 0.633–0.88), reflecting moderate discriminative performance; E) Variable importance in projection scores identifying the most influential biomarkers contributing to group separation.

The model demonstrated moderate goodness-of-fit (R²Y = 0.393), indicating that approximately 39% of the variance in group classification was explained by the model. However, the predictive ability was limited (Q² = 0.113, 7-fold cross-validation), suggesting modest generalizability to unseen data (fig.8-B).

Permutation testing confirmed that the model performed significantly better than chance (empirical p < 0.01), indicating that the observed class discrimination was not due to random variation (fig.8-C). Additionally, receiver operating characteristic (ROC) analysis yielded an area under the curve (AUC) of 0.758 (95% CI: 0.633–0.88), indicating moderate discriminatory performance (fig.8-D). VIP scores identified several key biomarkers contributing to group discrimination. The most influential features included: CD4⁺CD62L⁺CD45RA⁻, CD4⁺CD45RA⁺CD62L⁺, CD69⁺CD38⁺ on M1 and M2 monocytes, DC%, CD95 expression on CD4⁺ T cells (fig.8-E).

## 3. Discussion

The comprehensive phenotypic analysis of monocyte, dendritic and T-cell subsets of LC and CFS patients with different severity of depression and anxiety symptoms, revealed significant differences in the activation, differentiation and functional status in comparison to healthy controls, as well as between the two pathological states.

Our finding of progressive M2 dominance (HC < CFS < LC) with reciprocal M1 reduction aligns with emerging evidence of immune exhaustion and altered macrophage activation in both conditions [42]. The significantly increased M2 percentage in LC, combined with elevated M2+CD80 expression, suggests a paradoxical state where anti-inflammatory M2 cells express costimulatory molecules typically associated with immune activation. This may reflect the persistent low-grade inflammation and monocyte activation documented in LC, where monocytes exhibit mixed phenotypes with both pro-inflammatory and regulatory features [43].

In contrast, CFS patients in our cohort showed intermediate M2 levels between healthy controls and LC, suggesting a distinct immunopathological trajectory. This finding is supported by recent single-cell RNA sequencing data from Sun et al., who observed a significant reduction in total monocyte numbers in ME/CFS patients, along with enhanced cell-cell communication networks converging on monocytes [44]. Furthermore, Zheng et al. identified monocytic dysregulation as a mechanism of fatigue in ME/CFS, with upregulation of inflammatory targets (IL1β, CCL2, TLR2, STAT1, IFIH1) predominantly enriched in classical monocytes [45]. This may suggest that while CFS exhibits monocyte activation, the pattern differs fundamentally from the M2-dominant, profibrotic phenotype characteristic of LC.

The concept of "trained immunity" may partially explain the persistent M2 polarization in LC. Gu et al. demonstrated that severe COVID-19 induces extensive epigenetic reprogramming in hematopoietic stem and progenitor cells (HSPCs), leading to durable alterations in monocyte phenotypes that persist despite their short lifespan [46]. IL-6 appears pivotal for imprinting these epigenetic modifications during acute infection, potentially explaining the sustained M2 phenotype and elevated inflammatory markers observed months after initial SARS-CoV-2 infection [46]. This mechanism may be less prominent in CFS, where the initiating infectious triggers are often more remote in time, and diverse, potentially resulting in different patterns of HSPC reprogramming.

Our finding of significantly elevated CD80 expression on both M1- like and M2- like monocytes in LC, in contrast to decreased CD80 in CFS, represents a critical distinction between these conditions. CD80 (B7-1) is a key costimulatory molecule that, together with CD86 (B7-2), provides essential signals for T cell activation through CD28 and CTLA-4 binding [47]. The upregulation of CD80 on both M1 and M2 monocytes in LC suggests ongoing antigen presentation and immune activation, consistent with Hopkins et al.’s observation of sustained CD86 expression on monocytes and dendritic cells 6-7 months post-COVID-19 [48].

The paradoxical elevation of CD80 on M2 cells in LC is particularly intriguing. M2 macrophages are traditionally considered anti-inflammatory and involved in tissue repair, yet their expression of costimulatory molecules suggests a hybrid activation state. This may represent a "failed resolution phenotype" where anti-inflammatory M2 cells attempt to activate adaptive immunity but cannot effectively resolve inflammation. Pinto et al. demonstrated that CD86 expression on monocytes in chronic Chagas disease influences an immunomodulatory profile through CTLA-4 engagement and regulatory T cell activation [49]. A similar mechanism may operate in LC, where elevated M2+CD80 could engage CTLA-4 on T cells, contributing to T cell exhaustion while maintaining chronic low-grade inflammation.

In contrast, the significantly decreased M1+CD80 expression in CFS suggests profoundly impaired immune induction ability rather than persistent activation. This aligns with Eaton-Fitch et al.’s gene expression analysis showing downregulated interferon signalling and immunoglobulin genes in ME/CFS, indicating a state of immune suppression distinct from the persistent activation observed in LC [50]. The reduced costimulatory capacity of monocytes in CFS may contribute to the immunodeficiency features frequently reported in this population, including increased susceptibility to infections and impaired NK cell function [26,51–53].

The significantly decreased CD69+CD38 expression in LC, despite the high M2 numbers, provides compelling evidence for immune exhaustion in the monocyte compartment. CD69 is an early activation marker rapidly upregulated upon immune cell stimulation, while CD38 is a multifunctional ectoenzyme involved in calcium signalling and NAD+ metabolism [54]. The co-expression of these markers typically indicates recent activation, and their reduction in LC M2 cells might suggests either anergy or exhaustion despite other indicators of an ongoing low-grade inflammation.

The above finding contrasts sharply with the elevated DC+CD38+CD69 expression in moderate LC. This compartment-specific activation pattern suggests that while dendritic cells remain activated and capable of antigen presentation, monocytes/macrophages have entered an exhausted or tolerized state. Aid et al. recently demonstrated that LC is characterized by persistent activation of proinflammatory and immune exhaustion pathways, including upregulation of JAK-STAT, IL-6, complement, and T cell exhaustion pathways [55]. Our data extends these findings to the myeloid compartment, raising the question whether immune exhaustion is not limited to T cells but affects multiple immune cell lineages.

The reduction in CD38 expression may have additional metabolic implications. CD38 is a major NAD+ consumer, and its activity has been linked to cellular senescence and metabolic dysfunction [54]. Reduced CD38 expression on M2 cells in LC could represent an adaptive response to preserve NAD+ pools in the context of chronic inflammation and metabolic stress. However, this may come at the cost of impaired calcium signaling and immune cell function, contributing to the persistent symptoms, characteristic of LC.

Our finding of decreased M1+CD197 (CCR7) in both pathological states, but more important in CFS represents a critical distinction in immune cell trafficking capacity between these conditions. CCR7 is essential for lymph node homing of T cells and dendritic cells, controlling their migration to areas where T cell priming and adaptive immune response initiation occur [56–58] The progressive reduction in CCR7 expression suggests increasingly impaired immune cell trafficking, and immune response induction, most severe in CFS.

The intermediate CCR7 expression in LC may reflect a partial recovery of the migratory capacity compared to CFS, but still represents significant impairment compared to healthy controls. Scott et al. demonstrated that LC patients with unresolved lung injury exhibit high monocyte expression of CXCR6 and preferential migration toward CXCL16, which is abundantly expressed in the lung [59] This suggests a shift in chemokine receptor expression patterns, with reduced CCR7-mediated lymph node homing and increased tissue-specific chemokine receptor expression. Such altered trafficking patterns could contribute to persistent tissue inflammation while impairing systemic immune surveillance.

Recent work by Chen et al. provides additional mechanistic insight, demonstrating that during inflammation, CCL21 is downregulated in lymph node high endothelial venules, and lymphocytes become dependent on oxysterols and the receptor EBI2 for inflamed lymph node entry [60]. This inflammatory switch in chemoattractant requirements may explain how immune cell trafficking is maintained despite reduced CCR7 expression in LC. However, in CFS, where CCR7 expression is most profoundly reduced, this compensatory mechanism may be insufficient, contributing to more severe immune trafficking defects.

The correlation between reduced CCR7 expression and disease severity is supported by genome-wide association studies showing strong associations between the CCL21/CCR7 axis and disease severity in multiple autoimmune conditions, including rheumatoid arthritis, Sjögren’s syndrome, and systemic lupus erythematosus [61] Therapeutic strategies targeting the CCR7 network have shown promise in regulating dendritic cell migration and managing inflammatory diseases, which raises the question whether CCR7 modulation could represent a potential therapeutic target in both CFS and LC [[62]

The elevated DC numbers in LC may reflect an ongoing process of antigen presentation and immune activation. Guerrera et al. identified an immunological signature of LC in some patients characterized by reduced plasmacytoid and conventional dendritic cell subsets, but with those remaining showing heightened activation [63]. This apparent contradiction may reflect heterogeneity within the LC population, with some patients showing DC depletion (similar to acute COVID-19) while others show DC expansion and activation. Our cohort appears to represent the latter phenotype, characterized by DC expansion and elevated costimulatory molecule expression.

In CFS, the intermediate DC numbers suggest a different trajectory of immune dysregulation. Sun et al.’s single-cell RNA sequencing study reported significant reductions in conventional and plasmacytoid dendritic cells in ME/CFS patients, while our flow cytometry data show intermediate levels between healthy controls and long COVID [44]. This discrepancy may reflect differences in patient selection, disease duration, or methodological approaches. However, both studies agree that DC alterations are a prominent feature of CFS immunopathology.

Our results show that moderate CFS and long COVID have significantly higher CD4+CD62L+CD45-RA- cells and lower CD4+CD45RA+CD62L- cells provide important insights into T cell dynamics in these conditions. This pattern may suggest a shift from naïve (CD45RA+) to central memory (CD62L+CD45RA-) phenotypes, representing ongoing immune activation and differentiation. The reduction in naïve T cells is consistent with T cell exhaustion and depletion of the naïve T cell pool, which may contribute to impaired responses to new antigens and ongoing immune dysregulation.

Wiech et al. demonstrated that T cell remodelling during LC depends on the severity of initial SARS-CoV-2 infection, with severe convalescents showing polarization toward exhausted/senescent CD4+ and CD8+ T cells, significant decrease in naïve cell populations, and increased terminal effector cells expressing CD57 [64]. Our data extend these findings to moderate disease, suggesting that naïve T cell depletion occurs across the severity spectrum but may be most pronounced in moderate cases. This could reflect a threshold effect, where moderate disease represents sufficient immune activation to drive T cell differentiation without the profound exhaustion observed in severe cases.

The increased CD4+CD62L+CD45-RA- cells in moderate disease may represent central memory T cells that have differentiated from naïve precursors. CD62L (L-selectin) is expressed on both naïve and central memory T cells and is essential for lymph node homing [27,65]. The expansion of CD62L+ cells despite reduced CD45RA+ naïve cells suggests active T cell differentiation and memory formation. However, this may come at the cost of depleting the naïve T cell pool, potentially impairing long-term immune competence.

Berentschot et al. reported that LC is characterized by T-lymphocyte senescence, with increased exhausted CD8+ T-lymphocytes expressing markers of terminal differentiation [42]. The severity of fatigue correlated with increases in intermediate and non-classical monocytes and higher CD8+ T-lymphocyte counts, suggesting that T cell and monocyte activation are linked in LC pathophysiology [42] Our finding of reduced naïve T cells in moderate disease supports this model, suggesting that ongoing immune activation drives T cell differentiation and exhaustion even in patients with moderate symptom severity.

The negative correlation between CD4+CD62L+CD45RA+ naïve/central memory cells and CD4+CD95+ apoptosis-prone cells in LC suggests that naïve T cell depletion is associated with increased T cell apoptosis. CD95 (Fas) is a death receptor that mediates activation-induced cell death, and its upregulation on T cells indicates susceptibility to apoptosis [64]. This mechanism may contribute to the progressive depletion of naïve T cells observed in moderate and severe LC, potentially explaining the long-term immune dysfunction characteristic of this condition.

The extensive correlation networks observed in both CFS and LC, with notably more complex patterns in LC, provide insights into the integrated nature of immune dysregulation in these conditions. In LC, the strong positive correlations between DC total numbers and M2-like populations, along with extensive correlations between CD80 expression across myeloid cell types, suggest coordinated activation of the myeloid compartment. This integration may reflect common upstream signals, such as persistent viral antigens, inflammatory cytokines, or metabolic factors that simultaneously affect multiple immune cell lineages.

The negative correlations between total M1/M2-like cell numbers and their activation markers (HLA-DR, CD80) in LC suggest a homeostatic mechanism where increased cell numbers are associated with decreased per-cell activation. This pattern may represent an attempt to maintain overall immune function while limiting excessive inflammation. However, this compensation appears insufficient, as LC patients continue to experience persistent symptoms and immune dysregulation.

In CFS, the correlation network reveals different patterns of immune dysregulation. The negative correlation between total DC and total M1 cells may reflect competitive differentiation pathways or reciprocal regulation between these cell types. The negative correlations between CD80/CD197 and CD69+CD38 in M2 cells suggest that activated cells have reduced costimulatory capacity and migratory potential, potentially contributing to impaired immune responses. These patterns differ fundamentally from LC, suggesting distinct immunopathological mechanisms.

The correlation between CD8+CD62L+CD45RA+ and CD4+CD62L+CD45RA+ cells in CFS, and the negative correlation between CD4+CD62L+CD45RA+ and CD4+CD95+ in LC, highlight different patterns of T cell regulation. In CFS, naïve/central memory CD4+ and CD8+ T cells appear co-ordinately regulated, suggesting T cell homeostasis in CFS is less affected in comparison to LC. In LC, the inverse relationship between naïve/central memory cells and apoptosis-prone cells suggests that T cell depletion is driven by activation-induced cell death, a mechanism consistent with chronic antigen exposure and immune exhaustion [55,64].

The positive association between T-cell differentiation markers and anxiety, depression, and fatigue is well-supported by recent evidence. Studies demonstrate that ME/CFS patients exhibit altered T-cell metabolism with reduced glycolysis and mitochondrial dysfunction, particularly in CD8+ T cells [66]. Severe ME/CFS is characterized by higher proportions of activated lymphocytes (CD69+, CD38+) and increased pro-inflammatory cytokine production (IFN-γ, TNF, IL-17), consistent with prolonged non-specific inflammation [67]. Long COVID with ME/CFS shows marked T-cell exhaustion, expansion of effector T cells, and depletion of naïve CD4+ and CD8+ T cells [16]. These alterations correlate with fatigue severity and cognitive impairment [42,68].

The T-cell differentiation axis likely reflects chronic immune activation and exhaustion, a state where persistent antigen exposure or viral reactivation drives T cells toward terminal differentiation [50,69]. This process is associated with increased inflammatory cytokines (IL-1, TNF-α) that directly correlate with fatigue, mood symptoms, and autonomic dysfunction [70,71]. The finding that this axis predicts symptom severity across multiple domains supports a model where adaptive immune dysregulation is a central driver of the post-viral syndrome phenotype.

The inverse relationship between monocyte HLA expression/antigen-presenting activity and psychiatric symptoms is indeed counterintuitive but has mechanistic precedent. While increased monocyte activation (intermediate and non-classical monocytes, CCL2/CCL7 expression) correlates with greater fatigue severity in Long COVID, the specific pattern of HLA-DR expression may reflect different functional states [42]. Recent single-cell analyses reveal that monocytes in Long COVID exhibit transcriptional skewing with reduced phagocytosis-associated genes and increased pro-inflammatory cytokine genes, suggesting functional heterogeneity within the innate compartment [16,43].

The protective association observed may reflect several mechanisms. First, adequate HLA-mediated antigen presentation is critical for viral clearance; weak HLA-antigen binding to viral antigens is associated with ME/CFS susceptibility, potentially allowing viral persistence [72]. Second, certain monocyte activation states may represent compensatory anti-inflammatory responses rather than pathogenic inflammation. Third, the timing and phase of immune activation matters—early robust innate responses may prevent progression to chronic adaptive dysregulation [73].

## 4. Materials and Methods

### 4.1. Patients’ Characteristics

This study was conducted and reported following STROBE guidelines for observational research. The study included a total of 207 participants, divided into 3 subgroups: (i) CFS group (n = 103), consisting of 28 males (43.96 ± 13.33 y.o.) and 75 females (46.75 ± 12.36 y.o.); (ii) long COVID (LC) group (n = 63), consisting of 17 males (42.88 ± 10.30 y.o.) and 46 females (49.96 ± 11.9 y.o.); and (iii) healthy control (HC) group (n = 41), consisting of 9 males (28.69 ± 10.17 y.o.) and 32 females (36.09 ± 15.5 y.o.).

### 4.2. Inclusion Criteria

CFS patients were included in the study according to the diagnostic criteria proposed by the Institute of Medicine (IOM) in 2015, based on the following three symptoms: 1. Significant reduction or limitation in the ability to perform pre-illness levels of daily activities that persists for more than 6 months, accompanied by fatigue that is often profound, of recent or definite (not lifelong) onset, and is not substantially relieved by rest. 2. Post-exertional malaise. 3. Unrefreshing sleep. At least one of the following is also required—cognitive impairment or orthostatic intolerance [29]. LC group participants were categorized, based on a positive SARS-CoV-2 PCR test and persistence of symptoms (fatigue/post-exertional malaise, dyspnea, joint pain, cognitive impairment, sleep disturbances, cardiovascular and gastrointestinal symptoms) that cannot be explained by other conditions for more than 4 months after recovery [30,31]. Healthy subjects were enrolled based on the absence of an active SARS-CoV-2 infection, confirmed by positive PCR/anti-SARS-CoV-2 IgG, and absence of: LC/CFS symptoms; systemic autoimmune diseases, administration of immunostimulatory or immunosuppressive drugs, history of chronic disease (including diabetes, impaired renal or hepatic function), hospitalization or viral/bacterial infection in the last 2 months preceding the study, abnormal complete blood count and/or biochemical status, increased C-reactive protein, chronic HIV infection, alcohol addiction and other dependencies.

### 4.3. Exclusion Criteria

We excluded pregnant and breastfeeding women. Subjects who suffered from psychiatric disorders such as major depressive disorder, generalized anxiety disorder, bipolar disorder, panic disorder, schizo-affective disorder, schizophrenia, psycho-organic syndrome, autism spectrum disorder, and substance use disorders (except tobacco use disorder (TUD), in attempt to avoid excessive exclusion) were not included in the study. We also excluded patients with cancer, systemic autoimmune diseases such as rheumatoid arthritis, diabetes mellitus, psoriasis, inflammatory bowel disease, neurodegenerative diseases (e.g., multiple sclerosis, Parkinson’s or Alzheimer’s disease) and stroke, as well as those with renal and liver disease. Medication including antidepressants (SSRIs), low-dose naltrexone, antihistamines, corticosteroids, beta-blockers, and antivirals in the last 2 months preceding the study was an exclusion criterion for both patients groups.

All participants signed a written consent. The study was conducted in accordance with the Declaration of Helsinki, and approved by the Scientific Ethics Committee at the Medical University of Plovdiv, Protocol No. 3/30 May 2023, for studies involving humans.

### 4.4. Clinical Examination

The participants’ health status, including their physical, mental, and behavioral well-being, were assessed by an experienced team of immunology specialists using a methodical interview. Sociodemographic, clinical, and psychological details were gathered through semi-structured interviews. Hamilton depression rating scale (HDRS) and the Hamilton anxiety rating scale (HAMA) were used to measure the degree of depression and anxiety, respectively. For this study, no physical symptoms from HAMD-17 (pure HAMD, pHAMD) and HAMA-14 (pure HAMA, pHAMA) were considered to calculate only the scores for pure depression and anxiety. Using HAMD, we created two subdomains: physiosomatic HAMD, which included somatic anxiety, gastrointestinal (GIS) anxiety, genitourinary anxiety, and hypochondriasis; and pure depression HAMD: depressed mood, guilt feelings, suicidal thoughts, and loss of interest. The HAMA was also divided into two subdomains: physiosomatic pHAMA—somatic sensory, cardiovascular, genitourinary, and autonomic symptoms, as well as GIS; pure anxiety HAMA—anxious mood, tension, fears, anxiety, and anxious behavior during the interview [11]. Additionally, a pure FibroFatigue-12 (pFF) score was calculated by adding the following items to the pFF scale: muscular pain, muscle tension, fatigue, autonomous symptoms, GIS, headache, and flu-like malaise [11,32–34]. qSUM was also evaluated, representing the total score from the questionnaires. Scores on the pHAMA-14, pHAMD-17, and pFF-12 were interpreted using established severity thresholds [32,35–40]. To capture the overall burden of neuropsychiatric and physiosomatic symptoms, a composite score was constructed by integrating results from the pHAMD, pHAMA, and pFF questionnaires. The established severity thresholds for each instrument were used as reference points, and a combined threshold was defined to reflect the presence of clinically relevant symptomatology across domains. This approach was adopted to provide a more comprehensive assessment of symptom burden, given the overlap between affective and somatic symptoms in the studied population. As no validated composite index exists for these instruments, this metric should be considered exploratory. This approach was chosen to account for the multidimensional nature of the condition, where fatigue, anxiety, and depressive symptoms frequently co-occur and may not be adequately captured by a single scale.

### 4.5. Sample Collection and Processing

Whole peripheral blood (6 mL) was collected into sodium heparinized collection tubes between 9 a.m. and 12 a.m. Peripheral mononuclear cells were isolated by gradient centrifugation as previously described [41], cryopreserved in fetal bovine serum (FBS; Corning) with 10% dimethyl sulfoxide (DMSO; Sigma Aldrich) and 60% RPMI 1640 media (Sigma Aldrich) and stored in liquid N2 until analysis. After thawing, cells were treated with 1ug/ml DNAse I (Roche Diagnostics, cat # 10104159001) and left for 2h at RT in 10% RPMI before staining for flow cytometry analysis.

### 4.6. Multicolor Phenotyping Analysis of Surface Markers

Multiparameter flow cytometry was used on peripheral blood mononuclear cells(PBMCs) to identify specific subpopulation of monocytes, T cells and dendritic cells. A 12-parameter flow cytometry panel was designed including the following surface markers: CD4-BUV496 (clone SK3, cat #612937), CD3-BUV-395 (clone UCHT1, cat #563546), CD80-BV786 (clone SK1, cat #344732 ), CD163-BV711 (GHI/61, cat #333630), CD69+CD38-BV650 (clone FN50/HB-7, cat #310934/356620), CD45RA-BV605 (clone HI100, cat #304134), CD8-BV510 (clone SK1, cat #344732), CD14-BV421 (clone M5E2, cat #301830), CD27-AlexaFluor700 (clone M-T271, cat #356416), CD206-APC (clone 15-2, cat #321110), CD62L-PE-Cy7 (clone DREG-56, cat #304822), CD197-PE-Cy5.5 (clone G043H7, cat #353220), and anti-HLA-DR-PE (clone L243, cat #307606). Samples were stained by adding pre-titrated concentrations of directly conjugated antibodies suspended in staining buffer (cat# 342417, BD, San Jose, CA, US), followed by 15 min incubation at room temperature in the dark, and repeated washing with PBS (Corning, cat # 21-031-CV). After preparation, samples were acquired immediately on FACS Aria II (BD Biosciences) and analysed with FACS Diva v. 9.0.1 (BD Biosciences, San Jose, CA, USA) and FlowJo software v.10.8.1 (FlowJo, LLC).

Gating strategy: singlets were isolated on a FSC-H vs. FSC-A plot, lymphocytes were defined on a FSC vs. SSC plot, and T cells were identified as CD3+ events. Monocytes and dendritic cells were gated initially on FSC vs. SSC plot and further identified as CD14+CD3- and CD3-CD14-HLA-DR+ events. M2-like monocytes were differentiated from M1-like based on the high expression of CD206, confirmed with high CD163 expression (Suppl.Fig.1). Results for CD markers with continuous expression are presented as median fluorescence intensity (MFI). Fluorescence Minus One (FMO) controls were used for setting gates on markers with continuous expression (CD206, CD80, HLA-DR, CCR7)

### 4.7. Statistical analysis

Statistical analyses were performed using IBM SPSS Statistics version 23.0 (IBM Corp., Armonk, NY, USA). Graphical representations were generated using GraphPad Prism version 8.0 (GraphPad Software, San Diego, CA, USA). Normality of data distribution was assessed using the Shapiro–Wilk test. As most variables did not show normal distribution, continuous data are presented as median with interquartile range (Q1–Q3). Group comparisons were performed using the Kruskal–Wallis test for continuous variables and the Chi-square test for categorical variables such as sex distribution. Effect sizes were calculated for non-parametric comparisons using rank-based measures to estimate the magnitude of observed differences. Associations between variables were assessed using the Spearman’s rank correlation. To account for differences in age between groups, a rank-transformed analysis of covariance (ANCOVA) was applied for selected parameters where age was identified as a potential confounding factor. Outcome variables were rank-transformed and entered into a general linear model with group as the main factor and age as the covariate. To control for multiple comparisons, false discovery rate (FDR) correction was applied using the Benjamini–Hochberg procedure. FDR correction was performed separately for each family of related statistical tests. Post hoc power analyses were conducted and demonstrated that all evaluated parameters had sufficient statistical power (≥0.80) to detect the observed differences. A p-value < 0.05 was considered statistically significant. Principal component analysis (PCA) was performed to explore multivariate relationships among measured biomarkers and clinical parameters. PCA was conducted for all subjects using standardized variables, and components with eigenvalues greater than 1 were retained according to the Kaiser criterion. PCA score plots were used to visualize clustering patterns and relationships among variables. Cases with missing values were excluded pairwise from the relevant analyses, and no data imputation was performed. Partial least squares discriminant analysis (PLS-DA) was conducted using MetaboAnalyst ver. 6.0. to assess group discrimination based on selected immune biomarkers. Data were log-transformed and auto-scaled prior to analysis. Model performance was evaluated using 7-fold cross-validation, and statistical significance was assessed using permutation testing. Variable importance in projection (VIP) scores were used to identify the most influential features.

## 5. Conclusions

This study demonstrates that chronic fatigue syndrome and long COVID, despite overlapping symptoms such as debilitating fatigue, display distinct immunophenotypic signatures that likely reflect different pathogenic mechanisms. Long COVID shows features of persistent immune activation characterized by progressive M2-like monocyte polarization, increased costimulatory molecule expression (particularly CD80), dendritic cell expansion, and immune exhaustion, suggesting a “failed resolution” state with possible ongoing antigen presentation. In contrast, CFS exhibits immune suppression with reduced M1-like costimulatory capacity, impaired CCR7-mediated immune cell trafficking, and more integrated systemic immune dysregulation, indicating defects in immune activation and migration rather than chronic overstimulation. Differences in T-cell composition, dendritic cell activity, and correlation network structure further support distinct immunological architectures between the diseases. Together, these findings raise the question whether these two conditions represent separate immunopathological entities, with implications for biomarker development, patient stratification, and the design of disease-specific therapeutic strategies targeting the underlying immune pathways, if validated in independent prospective cohorts.

## 6. Limitations

Several limitations of this study warrant consideration. The cross-sectional design limits assessing temporal dynamics of immune changes, and longitudinal studies are needed to determine whether the observed patterns represent stable disease states or evolving processes. The sample size, while adequate for identifying major immunophenotypic differences, may be insufficient to detect subtle variations or to fully characterize disease heterogeneity within CFS and long COVID populations with different disease severity. Phenotypic data would be more convincing if completed with functional assays. Additionally, our study is focused on peripheral blood immune cells, while tissue-resident immune populations may exhibit different patterns of activation and exhaustion.

## Data Availability

All data produced in the present work are contained in the manuscript

## Supplementary Materials

The following supporting information can be downloaded at: https://www.mdpi.com/article/doi/s1, Figure S1: Gating strategy

## Author Contributions

Conceptualization, S.P., H.T., and M.N.; methodology, S.P., Y.T.; validation, S.P., M.N., Y.T., and R.D.; formal analysis, S.P.; investigation, S.P., Y.T., R.D., M.N.; data curation, S.P.; writing—original draft preparation, S.P.; writing—review and editing, M.N., Y.T.,R.D., M.I., T.K., M.B., M.M. (Marianna Murdjeva),M.M. (Michael Maes) and H.T.; visualization, S.P.; supervision, M.N. and H.T.; All authors have read and agreed to the published version of the manuscript. **Funding**

“Strategic Research and Innovation Programme for the Development of the Medical University–Plovdiv (SRIPD-MUP)” Contract No BG-RRP-2.004-0007-C01”.

## Institutional Review Board Statement

The study was conducted in accordance with the Declaration of Helsinki and approved by the Scientific Ethics Committee at Medical University of Plovdiv, Protocol No. 3/30, 30 May 2023, for studies involving humans.

## Informed Consent Statement

Informed consent was obtained from all subjects involved in the study.

## Data Availability Statement

Data are contained within the article.

## Abbreviations

The following abbreviations are used in this manuscript:

ME/CFS: Myalgic encephalomyelitis/chronic fatigue syndrome
COVID: Coronavirus disease
PCA: Principle Component Analysis
DC: Dendritic cells
MAIT: Mucosal-associated invariant T cells
NK: Natural killer cells
HDRS: Hamilton depression rating scale
HAMA: Hamilton anxiety rating scale
TUD: Tobacco use disorder
HIV: Human immunodeficiency virus
PBMCs: peripheral blood mononuclear cells
MFI: Median fluorescence intensity
FSC: Forward scatter
SSC: Side scatter
FMO: Fluorescence minus one
ANCOVA: analysis of covariance
FDR: False discovery rate
PLS-DA: Partial least squares discriminant analysis
VIP: Variable importance in projection
LC: Long COVID-19
FF: Fibro fatigue
IL: interleukin
RNA: Ribonucleic acid
CD: Cluster of differentiation
CCR: CC chemokine receptor
CXCL: chemokine (C-X-C motif) ligand
IFN: interferon
TNF: Tumor necrosis factor
HLA: Human leukocyte antigen
CTLA: Cytotoxic T-lymphocyte associated protein

