## Supplementary figures and images for "Comprehensive Immunophenotyping of Monocytes and Dendritic Cells Suggests Distinct Pathophysiology in Chronic Fatigue Syndrome and Long COVID"

### Supplemental Figure 1

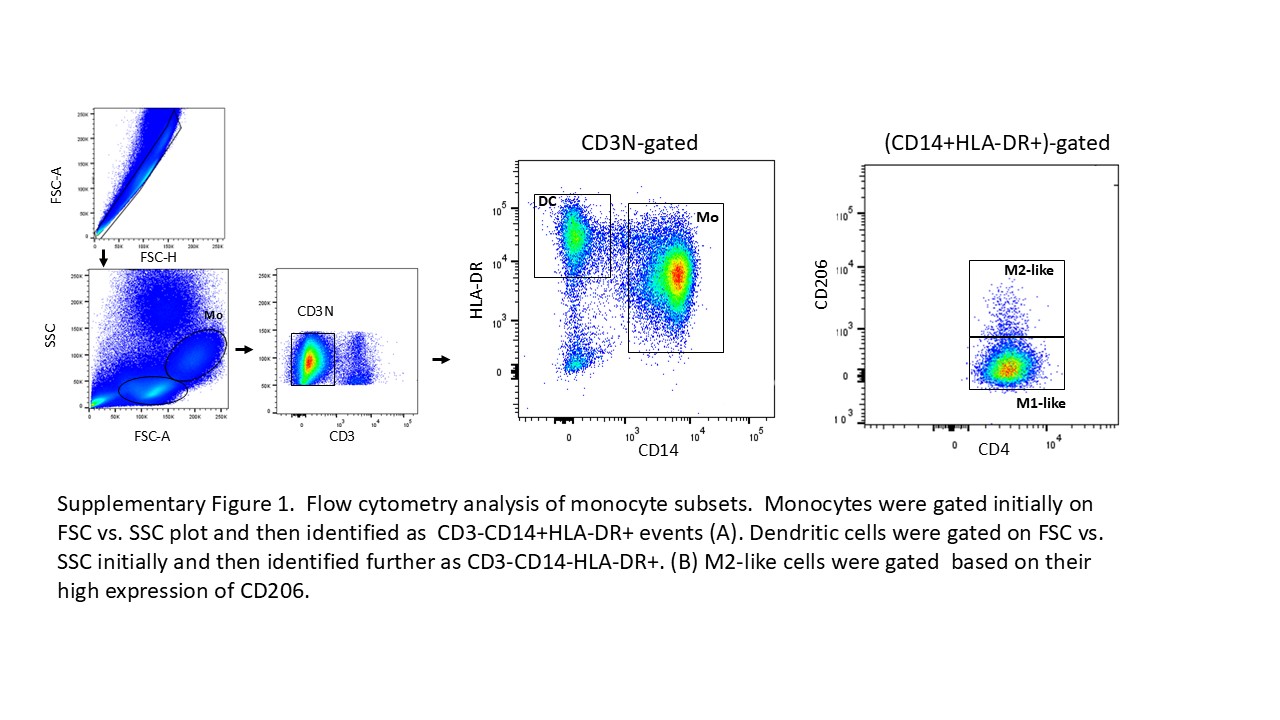
